# Assessing the usage of indirect motor pathways following a hemiparetic stroke

**DOI:** 10.1101/2020.12.23.20248683

**Authors:** Runfeng Tian, Julius P.A. Dewald, Yuan Yang

**Affiliations:** Stephenson School of Biomedical Engineering, University of Oklahoma, Tulsa, OK; Department of Physical Therapy and Human Movement Sciences, Feinberg School of Medicine and the Department of Biomedical Engineering, Northwestern University, Chicago, IL; Stephenson School of Biomedical Engineering, University of Oklahoma, Tulsa, OK and Department of Physical Therapy and Human Movement Sciences, Feinberg School of Medicine and the Department of Biomedical Engineering, Northwestern University, Chicago, IL

**Keywords:** brain-muscle connectivity, hemiparetic stroke, movement impairment, nonlinear analysis

## Abstract

A hallmark impairment in a hemiparetic stroke is a loss of independent joint control resulting in abnormal co-activation of shoulder abductor and elbow flexor muscles in their paretic arm, clinically known as the flexion synergy. The flexion synergy appears while generating shoulder abduction (SABD) torques as lifting the paretic arm. This likely be caused by an increased reliance on contralesional indirect motor pathways following damage to direct corticospinal projections. The assessment of functional connectivity between brain and muscle signals, i.e., brain-muscle connectivity (BMC), may provide insight into such changes to the usage of motor pathways. Our previous model simulation shows that multi-synaptic connections along the indirect motor pathway can generate nonlinear connectivity. We hypothesize that increased usage of indirect motor pathways (as increasing SABD load) will lead to an increase of nonlinear BMC. To test this hypothesis, we measured brain activity, muscle activity from shoulder abductors when stroke participants generate 20% and 40% of maximum SABD torque with their paretic arm. We computed both linear and nonlinear BMC between EEG and EMG. We found dominant nonlinear BMC at contralesional/ipsilateral hemisphere for stroke, whose magnitude increased with the SABD load. These results supported our hypothesis and indicated that nonlinear BMC could provide a quantitative indicator for determining the usage of indirect motor pathways following a hemiparetic stroke.

## I. Introduction

THE human motor system is a highly cooperative network comprised of different groups of neurons. Neural connectivity, i.e., the synchronization of neural activity across these groups, is key to the coordination among distant, but functionally related, neuronal groups during the control of movement [1]. After a hemiparetic stroke, damage to the brain increases reliance on indirect motor pathways resulting in motor impairments [2-4] and changes in neural connectivity [5]. A hallmark of impairments post hemiparetic stroke is a loss of independent joint control resulting in the abnormal coupling between shoulder abductor and elbow/wrist/finger flexor muscles, known as the flexion synergy [6]. The flexion synergy limits arm/hand function, like reaching and hand opening; and has also been reported to be linked to hyperactive stretch reflexes or spasticity [7, 8].

The flexion synergy appears while generating shoulder abduction (SABD) torques [9, 10] when lifting the weight of the arm or more. It is thought to be caused by progressive recruitment of contralesional indirect motor pathways via the brainstem as a function of SABD effort following a stroke-induced loss of ipsilesional corticospinal projections [2, 11, 12]. Thus, a neural connectivity measure that quantifies the recruitment of these indirect motor pathways would be crucial to evaluate post-stroke motor impairments. This measure also allows for the determination of the effect of new therapeutic interventions that aim to reduce such recruitment thus improving paretic arm function.

For decades, researchers investigating neural connectivity of motor pathways focused on linear (iso-frequency) neural coupling between brain (EEG) and muscle activity (EMG), i.e., EEG-EMG coherence [13], by assuming that motor commands from cortex to muscle are linearly transmitted [14]. This linear measurement primarily reflects neural connectivity of direct corticospinal tracts, and reduces at the ipsilesional hemisphere post hemiparetic stroke [5, 15]. However, no enhanced EEG-EMG coherence was found at the contralesional hemisphere during the flexion synergy expression[5, 15], indicating that this measurement may not allow examining the usage of contralesional indirect motor pathways. A plausible reason could be that the neural connectivity of indirect motor pathways is nonlinear [16, 17], showing neural coupling across different frequencies.

Recently, we developed a novel method, namely cross-spectral connectivity, to assess both linear and nonlinear connectivity that cannot be studied by using traditional connectivity methods such as linear coherence [18]. Our neural model simulation indicated that nonlinear connectivity is likely generated by the nonlinear behavior of synaptic connection and can be cumulatively enhanced across more synapses as in indirect motor pathways [19]. As shown in the simulation, multi-synaptic interactions of indirect motor pathways can lead to more dominant nonlinear connectivity between brain and muscle activity, in comparison to direct corticospinal tracts containing only one/two synapses [19].

In the present study, we investigated the nonlinear vs. linear connectivity between the brain and muscles in hemiparetic stroke during different levels of SABD tasks. We hypothesize that increased usage of indirect motor pathways while lifting the paretic arm, requiring SABD and causing the flexion synergy, will lead to a more dominant nonlinear connectivity between brain and muscle activity. To test this hypothesis, we measured scalp EEG, EMG from shoulder abductor muscles as well as flexion synergy elbow torques when eight stroke participants generate 20% and 40% of maximum SABD torque with their paretic arm. By testing the hypothesis, this proof-of-study will provide us with a verified quantitative measure to assess the change to the usage of motor pathways in hemiparetic stroke.

## II. Materials and Methods

### A. Participants and Data Collection

Eight individuals with chronic hemiparetic stroke (age: 64.4 ± 8.0 yrs.) and eight age-matched able-bodied participants (age: 60.1 ± 7.5 yrs.) were included in this proof-of-concept study. The participants were recruited with written informed consent and permission of the Northwestern University institutional review board. All participants with stroke were screened for inclusion by a licensed physical therapist. Inclusion criteria included 1) upper limb paresis, 2) ability to sit and lift the paretic arm without support, 3) an Upper Extremity Fugl Meyer Assessment (UE-FM) no greater than 40 out of 66, 4) more than one-year post-stroke, and 5) subcortical lesions not extending into sensorimotor cortices. Demographic information for each participant is provided in Table 1.

**TABLE I.**
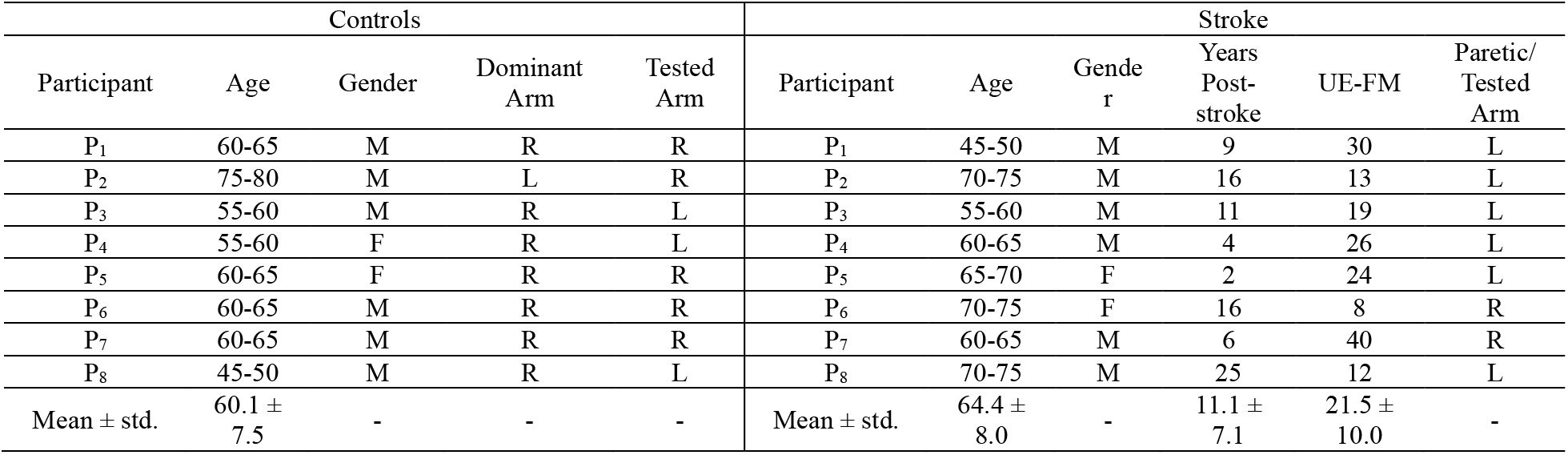
Participant Demographics

During the experiment, the participants were seated in a Biodex pedestal attached to a Biodex track (System 3 ProTM., Shirley, NY) with their trunk secured by belt restraints across the chest and abdomen. The tested arm was cast by fiberglass to line up the forearm, wrist, and hand and then rigidly attached to a customized beam connected to a JR3 6-DOF load cell (JR3, Woodland, CA). The tested upper limb was positioned with 85° shoulder abduction, 45° shoulder flexion, 90° elbow flexion. The load cell can measure the SABD and EF torques generated by the participant simultaneously. The medial epicondyle of humerus was aligned with the central axis for the rotation of the the load cell. The upper limber was positioned with the shoulder at 85° abduction, 45° flexion, and the elbow at 90° flexion angle.

At the beginning of the experiment, the participants were asked to lift the tested arm with their most effort. The average of three trials without large variance was considered as the maximum SABD torque of the participant. Then for each trial, the participants were asked to lift the tested arm against 20% (SABD20) or 40% (SABD40) of maximum voluntary torque (MVT) after receiving an auditory cue and hold for 10 seconds. Visual feedback was provided by a monitor in front of the participant. A red line would rise in proportion to the SABD torque, and the trial started when the target torque was reached. If the SABD torque fell off the 10% error range of the target torque, the participant would be asked to relax and restart the trial after enough rest. Participants performed 25 trials of each level task in total, with 5 trials of one level and shifted to 5 trials of another. Between trials, participants were asked to relax their arm fully for at least 1 minute. 32-channel EEG was recorded using active electrodes (Biosemi, Inc, Active II, Amsterdam, the Netherlands) mounted on a cap with the 10/20 system. The muscle activity at Intermediate Deltoid of the tested arm was recorded simultaneously by the differential of 2 Biosemi active electrodes with 1 cm inter-electronic distance. All data were sampled at 2048 Hz. The impedance of all electrodes was kept below 25 kΩ during the experiment.

### B. Data analysis

#### 1) Preprocessing

EEG and EMG data were preprocessed using EEGLAB. Raw EEG data were band-pass filtered between 1 and 100 Hz and notch filtered between 58 to 62 Hz (to remove line noise) with a zero-phase shift filter. Independent Component Analysis (ICA) algorithm was applied on filtered EEG to remove artifacts caused by movement and eye-blinks. Raw EMG data were band-pass filtered between 20 and 100 Hz and notch filtered between 58 to 62 Hz with a zero-phase shift filter, rectified, and normalized to the peak rectified EMG obtained during maximum SABD. Then all data were segmented into 1 s epochs with 250 ms overlapping. EEG signal was examined to remove bad channels and epochs with artifacts.

#### 2) Laterality Index of EEG power

The signal power of task-related brain activity at each EEG channel was estimated by the root-mean-square of the ensemble-averaged (over epochs) EEG. The Laterality Index was then be computed to indicate hemispheric dominance of brain activity: *LI* = (*CS* − *IS*)/(*CS* + *IS*), where CS is the sum of estimated signal power over the contralateral/ipsilesional sensorimotor areas, and IS is the sum over ipsilateral/contralesional sensorimotor areas. LI > 0 indicates the contralateral/ipsilesional dominance, LI < 0 the ipsilateral/contralesional dominance.

#### 3) EEG-EMG connectivity and N-L index

We computed the EEG-EMG connectivity (separating its nonlinear vs. linear parts) during shoulder abduction to investigate flexion synergy related neural connectivity by using our cross-spectral connectivity (CSC) method [18]. The CSC is a nonlinear extension of the classical (linear) coherence (used in the corticomuscular coherence), based on high-order spectra for distinguishably measuring nonlinear (cross-frequency) and linear (iso-frequency) connectivity between signals:

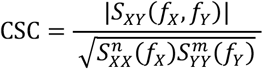

where *f*_*X*_ and *f*_*Y*_ are frequencies of EEG and EMG, *n* and *m* are integers following *n*: *m* = *f*_*X*_: *f*_*Y*_, and 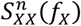 is the n-th order auto-spectra. The linear part of our results obtained by this method, when *n* = *m*, is comparable to the commonly used corticomuscular coherence. To compare the dominance of nonlinear vs. linear connectivity, we define the nonlinear-over-linear index (N-L Index): N-L Index = (SN-SL) / (SN+SL), where SL is the sum of linear connectivity over contralateral/ipsilesional sensorimotor areas, SN is the sum of nonlinear connectivity over ipsilateral/contralesional sensorimotor areas, N-L Index > 0 indicates greater nonlinear connectivity, N-L Index < 0 indicates greater linear connectivity. This definition is in line with our previous study for comparing linear and nonlinear connectivity in able-bodied individuals [20].

#### 4) Statically Analysis

We applied the Shapiro-Wild test [21] and Levene’s F-test [22] to verify the normality and homoscedasticity to allow parametric analysis. Student’s two-sample one-tailed paired t-test was applied to compare the group means for 20% SABD vs. 40% SABD levels within each group, and Student’s two-sample one-tailed t-test was applied to compare the group means of stroke vs. control at the same level of SABD. When the group is heteroscedastic and parametric analysis was not applicable, Welch’s two-sample one-tail (paired) t-test was applied. A significance level of 0.05 was used for all the tests in this paper.

## III. Results

Shown in all participants suffering from a hemiparetic stroke, the expression of the flexion synergy enhanced with increased SABD load (see Figure 1, paired t-test: 0.0003).

**Figure 1.**
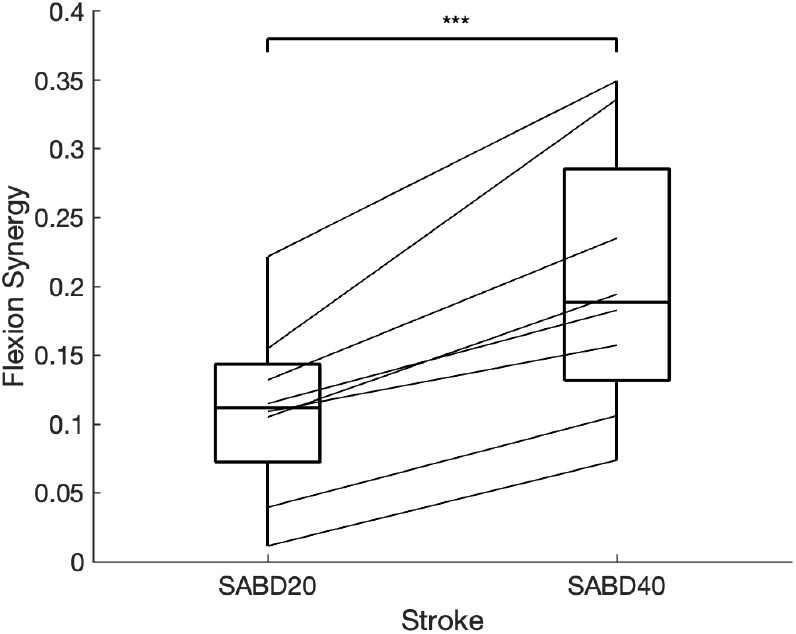
Flexion synergy of hemiparetic stroke subjects with different level of shoulder abduction. Flexion synergy is represented by the normalized involuntary elbow flexion during shoulder abduction tasks. On each box, the central mark indicates the median, and the bottom and top edges of the box indicate the 25th and 75th percentiles, respectively. Paired t-test was applied. * for p-value < 0.05, ** for p-value <0.01 and *** for p-value <0.001.

Participants suffering from a hemiparetic stroke show a lower Laterality Index (see Figure 2) in comparison to the able-bodied controls (two-sample t-test p-value< 0.00001 for both SABD20 and SABD40), indicating a more dominant ipsilateral brain activity in hemiparetic stroke during the SABD tasks. Meanwhile, a higher N-L index (see Figure 3) is shown in the stroke participants as compared to the able-bodied controls (two-sample t-test p-value < 0.00001 for both SABD20 and SABD40), indicating a more dominant nonlinear BMC in hemiparetic stroke. When the stroke participants were lifting their paretic arm more as increasing their SABD load from 20% to 40% MVT, both the laterality index and the N-L index significantly increase their values (paired t-test p-value = 0.0004 for Laterality Index and p-value = 0.0016 for N-L index). No such significant changes were shown in the able-bodied controls when they were increasing the SABD load with the same amount.

**Figure 2.**
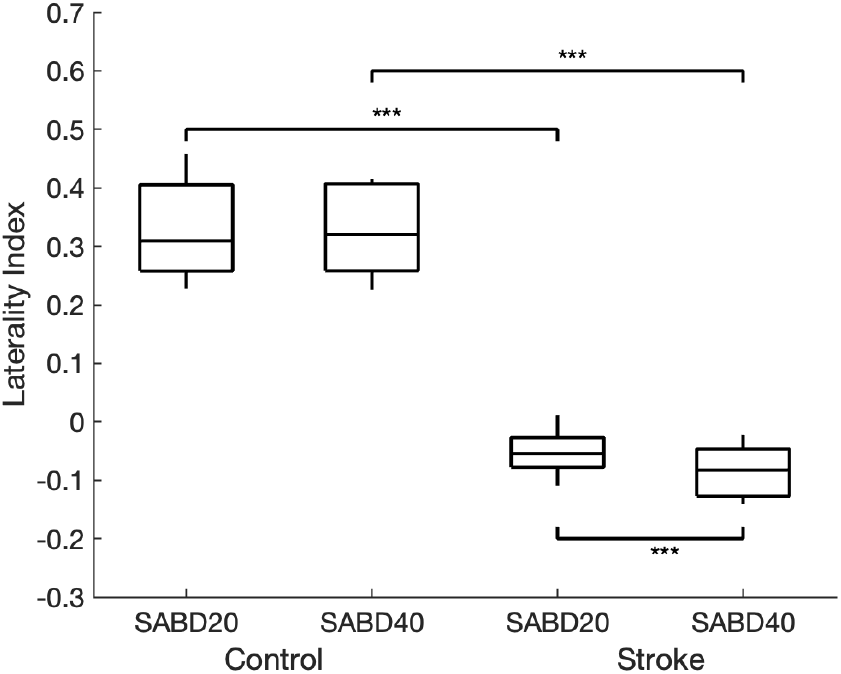
Laterality index for control and stroke subjects with different level of shoulder abduction. Two-sample t-test was applied across groups with same level of SABD, and paired t-test was applied among groups with different levels of SABD. * for p-value < 0.05, ** for p-value <0.01 and *** for p-value <0.001.

**Figure 3.**
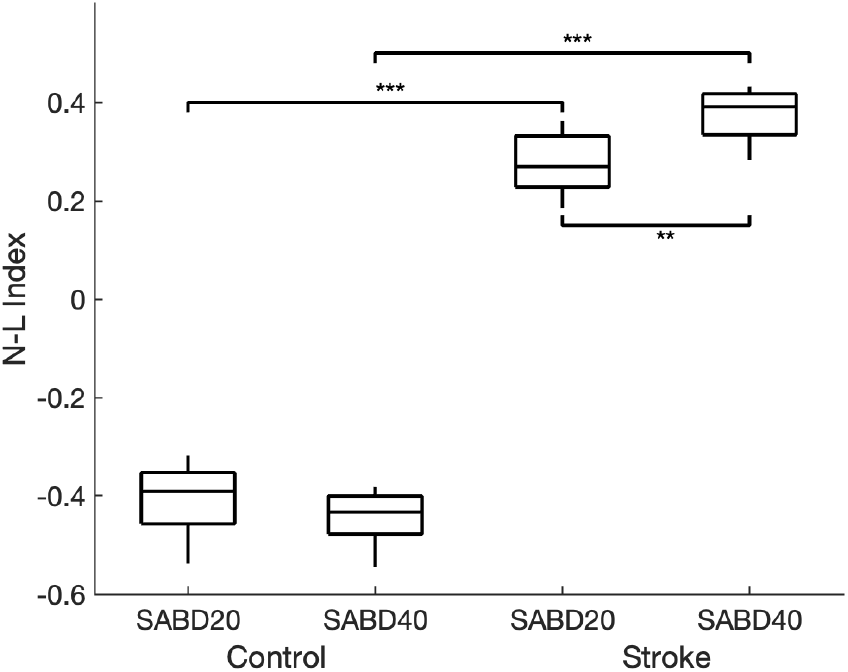
N-L index for control and stroke subjects with different level of shoulder abduction. Two-sample t-test was applied across groups with same level of SABD, and paired t-test was applied among groups with different levels of SABD. * for p-value < 0.05, ** for p-value <0.01 and *** for p-value <0.001.

## IV. DISCUSSION

Recruitment of contralesional indirect motor pathways post-stroke is thought to be the cause of the expression of flexion synergy in chronic hemiparetic stroke [2]. Quantitative measurement of the use of indirect motor pathway is essential to evaluate the motor impairments and guide the target intervention that aims to reduce the expression of flexion synergy. Traditional linear coherence was not capable to address the brain-muscle connectivity because of the accumulation of nonlinearity after the motor command passing through multiple synapses in the indirect motor pathways via the brain stem [19]. By applying our cross-spectral connectivity (CSC) method, we, for the first time, assessed both linear and nonlinear brain-muscle connectivity (BMC) at different SABD levels (20% vs. 40%) in chronic hemiparetic stroke, as compared to the healthy controls. We found that nonlinear connectivity is dominant as N-L index > 0 in hemiparetic stroke, indicating that stroke subjects tend to use more the indirect, multi-synaptic motor pathways, when they were performing the SABD tasks, due to the damage to their direct corticospinal tract. Furthermore, the laterality index < 0 indicates that the recruited indirect motor pathways are from the contralesional hemisphere (ipsilateral to the paretic arm). This result is in line with our recent imaging work using diffusion tensor imaging (DTI) demonstrated that damage to the corticospinal tract can lead to the increased structural integrity of indirect motor pathways (e.g., the medial reticulospinal tract) at the non-lesioned side [23] in chronic hemiparetic stroke.

The N-L index increases when the stroke subjects were performing a higher level SABD task (40%) while the laterality index decreases. This is likely because the use of contralesional indirect motor pathways increases with the SABD level since a higher level SABD task requires more effort of contralesional indirect motor pathways to compensate for the lost function of the direct corticospinal tract.

In healthy controls, the linear connectivity is dominant N-L index < 0 and it is mainly from the contralateral hemisphere as the laterality index > 0. This result is in line with previous studies showing the healthy controls are mainly using the direct, contralateral corticospinal tract for motor control of upper limb muscles [16, 17] and, therefore, showing the dominance of the contralateral, linear BMC [13, 20].

## V. CONCLUSION

The applied cross-spectral connectivity method allows the assessment of both linear and nonlinear brain-muscle connectivity that cannot be quantified by using conventional connectivity methods such as coherence which only assesses linear brain-muscle connectivity. The results in this study support our hypothesis that increased usage of indirect motor pathways leads to a more dominant nonlinear connectivity between brain and muscle activity as reflected by an increased N-L index. As such, this study, for the first time, provides a sensitive metric (i.e., N-L index) to quantitatively determine the usage of indirect motor pathways and its link to the expression of flexion synergy. The obtained knowledge and the proposed measures are essential for future studies examining how the recruitment of indirect motor pathways can be minimized by using more targeted interventions, such as physical [24] and pharmacological [25] methods we recently proposed to promote the use of remaining corticospinal resources from the lesioned hemisphere post hemiparetic stroke.

## Data Availability

The data that support the findings of this study are available from the corresponding author, Y. Yang, upon reasonable request.

## Acknowledgment

The authors would like to thank Mark Quinlan Cummings for his assistance in participant recruitment.

